# Between Patterns and Predictions: Interpretable Latent EEG Representations for Clinical Insights

**DOI:** 10.64898/2026.06.12.26355536

**Authors:** Laura Krumm, Robert Terziev, Dominik D. Kranz, M. Brandon Westover, Christian Meisel

## Abstract

Electroencephalography (EEG) captures rich brain dynamics, yet in clinical practice this complexity is often reduced to simplified summaries or categorical labels, limiting its interpretability for decision-making. We tested the hypothesis that a pretrained latent embedding framework, the Universal Map of EEG (UM-EEG), can preserve clinically meaningful structure across heterogeneous datasets and provide a generalizable representation of brain states. We applied UM-EEG, without retraining, to three independent cohorts spanning distinct clinical contexts: long-term EEG recordings from cardiac arrest patients (n = 576), subarachnoid hemorrhage (n = 100), and routine clinical EEG recordings containing physiological and pathological patterns (n = 141). EEG segments were projected into a shared 128-dimensional space anchored by expert-derived reference states, including wakefulness, sleep stages, ictal-interictal continuum activity, and burst suppression. Across datasets, favorable outcome or physiological recordings were consistently located closer to healthy reference states, whereas poor outcome and pathological recordings shifted toward pathological regions of the embedding space. Trajectory-derived geometric and temporal features discriminated outcome in cardiac arrest (ROC-AUC 0.83) and subarachnoid hemorrhage (ROC-AUC 0.76), and distinguished physiological from pathological routine EEGs (ROC-AUC 0.93). In routine EEG, similarity relationships derived from embedding trajectories correlated with those derived from structured clinical reports, indicating that the latent space recapitulates clinically relevant organization. These findings show that a fixed, semantically structured EEG embedding generalizes across etiologies and recording settings, enabling prognostic stratification and contextual interpretation while preserving the relational structure of brain states.

## Introduction

Clinical decision-making in neurology relies on integrating electrophysiological signals with clinical examination and complementary diagnostic tests.^1^ In practice, however, this high-dimensional information is often compressed into simplified summaries that capture only a fraction of the underlying dynamics. Electroencephalography (EEG) provides a direct window into brain activity, yet its interpretation remains time-consuming, expertise-dependent, and difficult to integrate systematically into clinical workflows.^2^ Consequently, much of the richness contained in EEG recordings remains underutilized in routine decision-making.

Machine learning (ML) has begun to address these limitations. However, most existing methods reduce complex, time-varying signals to discrete classifications^3^ or scalar probability scores. While such outputs are convenient, they discard much of the structure inherent in the data and provide limited insight into the relational structure of brain states. This contrasts with clinical reasoning, in which EEG patterns are interpreted by relating recordings to known physiological and pathological states within a broader context rather than assigning them to a single category. This mismatch between model outputs and clinical reasoning may contribute to the limited translation of ML methods into clinical practice.

An alternative paradigm is to use ML to represent, rather than collapse, complexity. In this paradigm, high-dimensional recordings are embedded into a continuous space in which distances reflect meaningful similarities between brain states.^4–6^ Such representations enable individual observations to be interpreted relative to reference patterns, allowing a more nuanced characterization of brain dynamics that aligns with expert interpretation. Rather than producing isolated predictions, the model defines a structured landscape in which both typical and atypical patterns can be contextualized.

We previously introduced this concept as a “Universal Map of EEG” (UM-EEG), a latent embedding space trained on expert-defined reference patterns spanning wakefulness, sleep stages, ictal-interictal continuum activity, and burst suppression. In this framework, EEG recordings are represented as trajectories within a continuous space, and new data can be projected into this shared representation without retraining. This enables recordings from different patients, time points and clinical contexts to be interpreted within a common reference system. However, it remains unclear whether such a representation generalizes across heterogeneous clinical settings and captures relationships that are meaningful for clinical use.

In this study, we test the hypothesis that a single pretrained embedding space captures a clinically meaningful structure of brain states across diverse settings and encodes information relevant for longitudinal characterization and prognosis. We apply the UM-EEG to independent datasets that differ in etiology, recording duration, and clinical context, including long-term EEG recordings from patients with subarachnoid hemorrhage (SAH)^7–9^ and cardiac arrest (CA)^10–12^, as well as routine clinical EEGs. We then evaluate whether this unified representation generalizes across these heterogeneous data and supports quantitative tracking of brain state evolution and prognostic stratification. More broadly, we assess whether embedding-based representations can shift EEG interpretation from discrete classification to structured, relational understanding for clinically interpretable decision support.

## Materials and Methods

### Datasets

This study builds on Krumm et al.,^4^ using the pre-trained UM-EEG model and continuous EEG data from patients diagnosed with a disorder of consciousness (DOC) following cardiac arrest (CA) without modification.^10–12^ Retrospective analysis of data for this project was conducted with waiver of informed consent under approved institutional review board protocols (BIDMC: 2022P000417; MGH: 2013P001024).

We evaluated the model’s generalizability by applying it to two novel datasets with the CA cohort (n=576) as a reference. The first dataset consisted of long-term EEG recordings from patients diagnosed with subarachnoid hemorrhage (SAH; n=100).^8,13^ The second comprised routine EEG (n=141) spanning both healthy and pathological patterns, extending the framework beyond neurointensive care long-term monitoring to a broader clinical context.

Neurological outcome was categorized as good or poor using dataset-specific scales: Cerebral Performance Category (CPC) for CA (CPC 1–2 vs. 5), Glasgow Outcome Scale (GOS) for SAH (GOS 4–5 vs. 1–3), and clinical assessment for routine EEG (physiological vs. pathological). For routine data, the EEG reports were additionally reviewed and categorized by a neurologist (R.T.). To systematically evaluate the unstructured text, we extracted 13 selected features based on the Standardized Computer-based Organized Reporting of EEG (SCORE) features^14^, a framework for standardized assessment and reporting of EEG, and institution-specific terminology (Table 1).

**Table 1:** Standardised questions for routine EEG report assessment to evaluate information provided in the EEG reports. Features were selected based on institution-specific terminology and SCORE.^14^.

| Item | Question | Answer |
| --- | --- | --- |
| 1 | What is the frequency of the background rhythm / PDR in Hz? | number |
| 2 | What is the amplitude in μV? | number |
| 3 | Was sleep recorded? | Yes / No |
| 4 | Is epileptic interictal activity present? | Yes / No |
| 5 | Are there periodic discharges? | Yes / No |
| 6 | Is there rhythmic delta activity? | Yes / No |
| 7 | Is there epileptic activity? | Yes / No |
| 8 | Is there burst-suppression? | Yes / No |
| 9 | Is the EEG physiological? | Yes / No |
| 10 | Is the EEG pathological? | Yes / No |
| 11 | Is there regional slowing? | Yes / No |
| 12 | Is there cerebral/brain dysfunction? | Yes / No |
| 13 | Is there interictal epileptic activity or signs of increased excitability? | Yes / No |

### Data Preprocessing

We applied a preprocessing pipeline that ensures consistency across the CA, SAH and routine EEG datasets with the UM-EEG framework. From the 113 SAH patients in the original dataset^7^, 100 were included in our analysis with 13 patients excluded due to corrupted or empty data files. SAH EEG data were stored as pre-segmented bipolar recordings, with metadata specifying segment start times. Pairs of consecutive 5-second segments were concatenated to form non-overlapping, consecutive, 10-second epochs. Segments that were not temporally consecutive were excluded. Across datasets artifact rejection was performed at the segment level: segments were excluded if any channel exhibited flat-line activity (near-zero variance), all-zero values, or extreme amplitudes exceeding ±1000 µV.

All SAH and routine EEG recordings were preprocessed in the following order: resampling to 200 Hz (requiring up sampling for the routine EEG dataset, originally recorded at 128 Hz), notch filtering to remove power-line noise (60 Hz for SAH, 50 Hz for routine EEG), bandpass filtering between 0.5 and 40 Hz, and re-referencing to the four bipolar channels used in UM-EEG (F3-C3, C3-O1, F4-C4, C4-O2). Routine EEG data were segmented into non-overlapping 10-second epochs. Each segment was transformed into a spectrogram using the multitaper method.^15^ Spectrograms were normalized using the global mean and standard deviation of the embedding model’s training corpus. Finally, spectrograms were passed to the UM-EEG’s pretrained neural network^4^ to obtain 128-dimensional (128D) latent embeddings. For each patient, this resulted in a time series of 128D embeddings (each data point corresponding to 10 seconds), referred to as a patient trajectory in UM-EEG. Segment timing was expressed as absolute recording time for routine EEG, or as time since cardiac arrest for CA and time since bleeding for SAH.

### Evaluation of Patient Trajectories in UM-EEG

We quantified patient trajectories across the three cohorts in UM-EEG in the following ways: first, we compared cohorts and their outcome groups as 2D visualizations in UM-EEG; second, we extract interpretable and clinically meaningful features from the 128D embeddings and correlated them with outcome; and third, we evaluated how well the categorized routine EEG report information corresponded to features extracted by UM-EEG.

#### Visualization of Trajectories in 2D UM-EEG Representation

For visualization, all 128D embeddings were projected into a shared two-dimensional space using the pretrained Uniform Manifold Approximation and Projection (UMAP)^16^ reducer from UM-EEG. Group-specific distributions (good and poor outcome for CA and SAH as well as physiological and pathological for routine EEG data) were converted into smooth density estimates using Gaussian kernel density estimation (KDE), computed over a common coordinate range to ensure direct visual comparability. Density values were normalized within each panel and visualized using a continuous colormap. Reference EEG embeddings were overlaid to compare density patterns to known UM-EEG phenotypes (Figure 1G). To quantify the patterns visible in the density plots, we computed the fraction of recording time each patient spent close to each of the 11 reference states and compared distributions between outcome groups using Mann-Whitney U tests.

**Figure 1.**
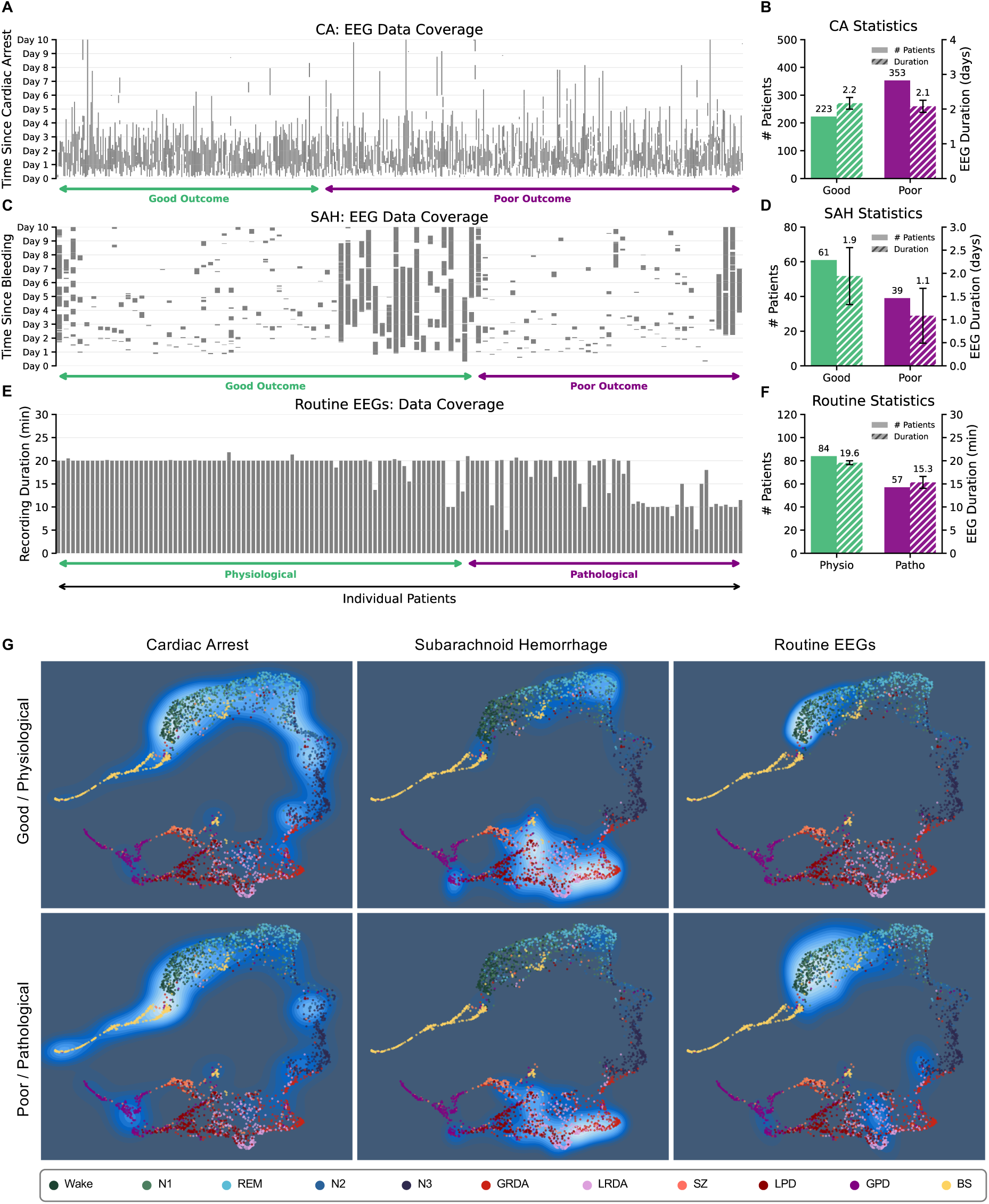
Dataset overview and 2D UM-EEG visualization. **A** EEG data coverage for individual CA patients up to 10 days after cardiac arrest; recordings exceeding 10 days were excluded due to sparse data. **B** Number of CA patients with good (green) and poor (purple) outcome and mean total EEG recording duration with 95% confidence interval (gaps excluded). **C** EEG data coverage for individual SAH patients up to 10 days after hemorrhage. **D** Number of SAH patients by outcome group and mean recording duration with 95% confidence interval. **E** Recording durations for individual routine EEG patients. **F** Number of routine EEG patients by physiological (green) and pathological (purple) label, and mean recording duration with 95% confidence interval. **G** 2D UMAP projection of all three cohorts into the shared UM-EEG embedding space, split by outcome group. Scatter points represent reference EEG embeddings with expert-assigned state labels. Patient embeddings are shown as Gaussian kernel density estimates computed over a common coordinate range to allow direct visual comparison across cohorts and outcome groups.

#### Quantification of Trajectories in UM-EEG

To quantify patient movement in the UM-EEG, each patient embedding was assigned to its most similar UM-EEG reference class using a support vector machine (SVM) trained on the UM-EEG test dataset. The classifier assigned each EEG segment to one of eleven reference states: physiological sleep stages (W (wake), REM (rapid eye movement sleep), non-REM sleep stages N1-N3)) or pathological patterns (SZ (seizures), GRDA (generalised rhythmic delta activity), LRDA (lateralised rhythmic delta activity), GPD (generalized periodic discharges), LPD (lateralised periodic discharges) and BS (burst suppression)). From the resulting label sequence for each patient, we extracted features across several categories: geometric features (mean angular distance from patient embeddings to each states’ centroid and to the combined healthy centroid), state occupancy (fraction of time and longest consecutive run in each state), entropy measures (entropy of label distributions), and temporal dynamics (self-transition probability), as listed in Table 2. This approach quantified each patient’s trajectory through the 128D embedding space, providing insight into how brain states move relative to reference states and enabling comparison across patients and cohorts.

**Table 2:**
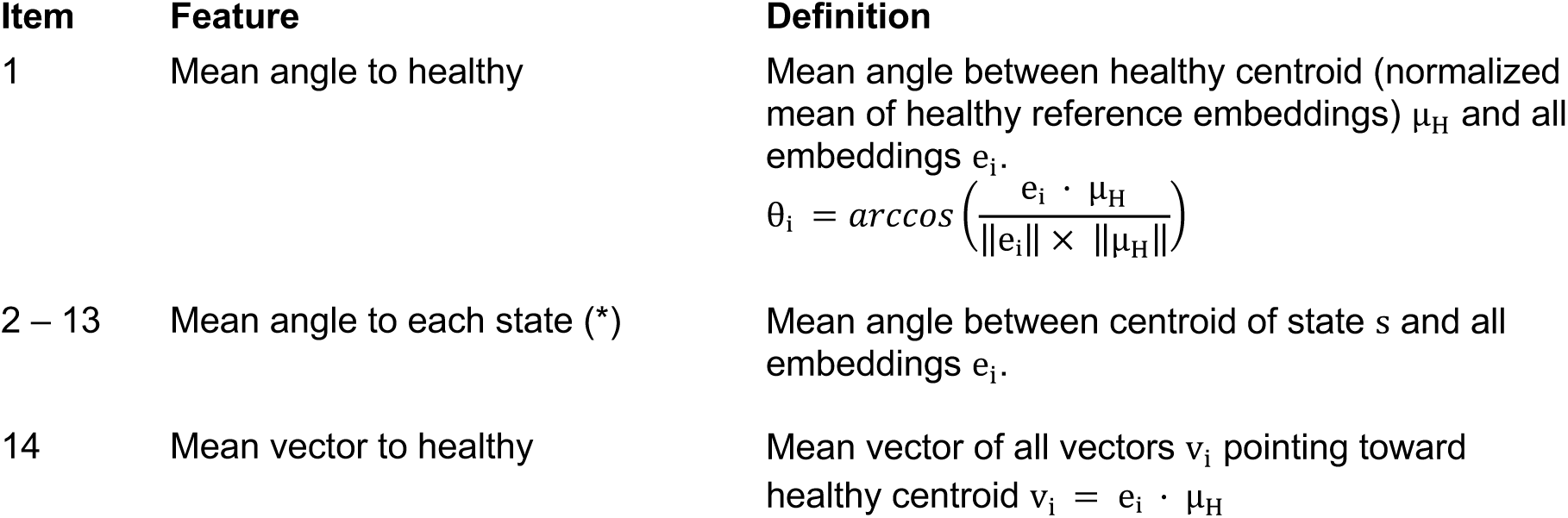
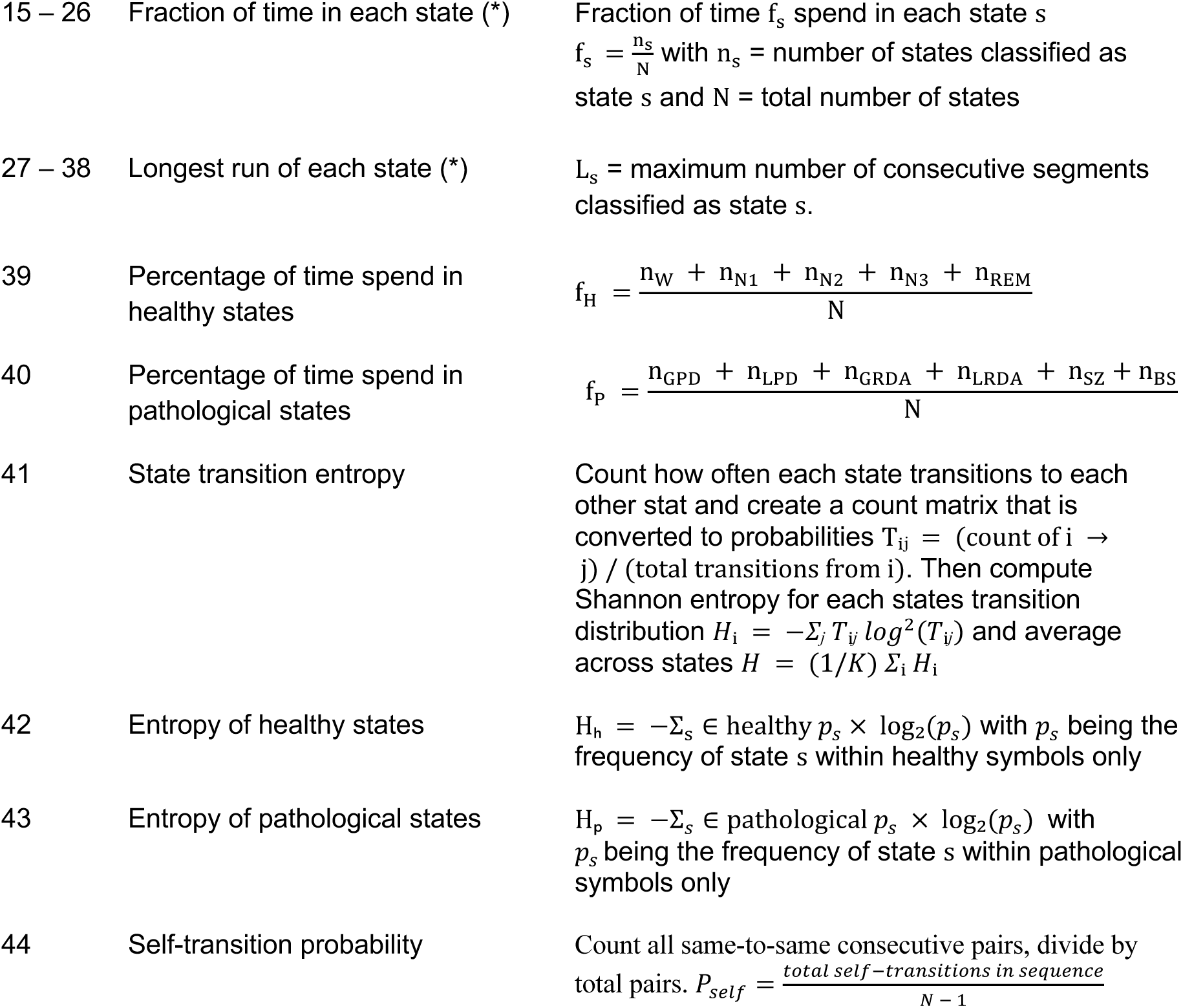
Trajectory features extracted from 128D embeddings. For all metrics above, the 128D embedding corresponding to each 10 second EEG segment is first classified by a linear SVM trained on UM-EEG reference embeddings. The class with maximum probability is assigned. The above features are then computed on the assigned label sequence per patient. (*) refers to the 11 UM-EEG reference states: wake (W), rapid eye movement sleep (REM), non-REM sleep stages N1-N3 (N1-N3), generalised and lateralised rhythmic delta activity (GRDA, LRDA), generalised and lateralised period discharges (GPD, LPD), seizures (SZ) and burst suppression (BS).

Optimal three-feature combinations that best distinguished good from poor outcome (CA and SAH) or physiological from pathological recordings (routine EEG) were identified using linear discriminant analysis (LDA) applied to all possible feature combinations. A hyperplane was fitted in this three-dimensional space, and receiver operating characteristic (ROC) curves with corresponding area under the curve (AUC) values were computed. 95% confidence intervals were estimated via bootstrap resampling with 50 iterations (Figure 2A, B, D, E, G, H).

**Figure 2.**
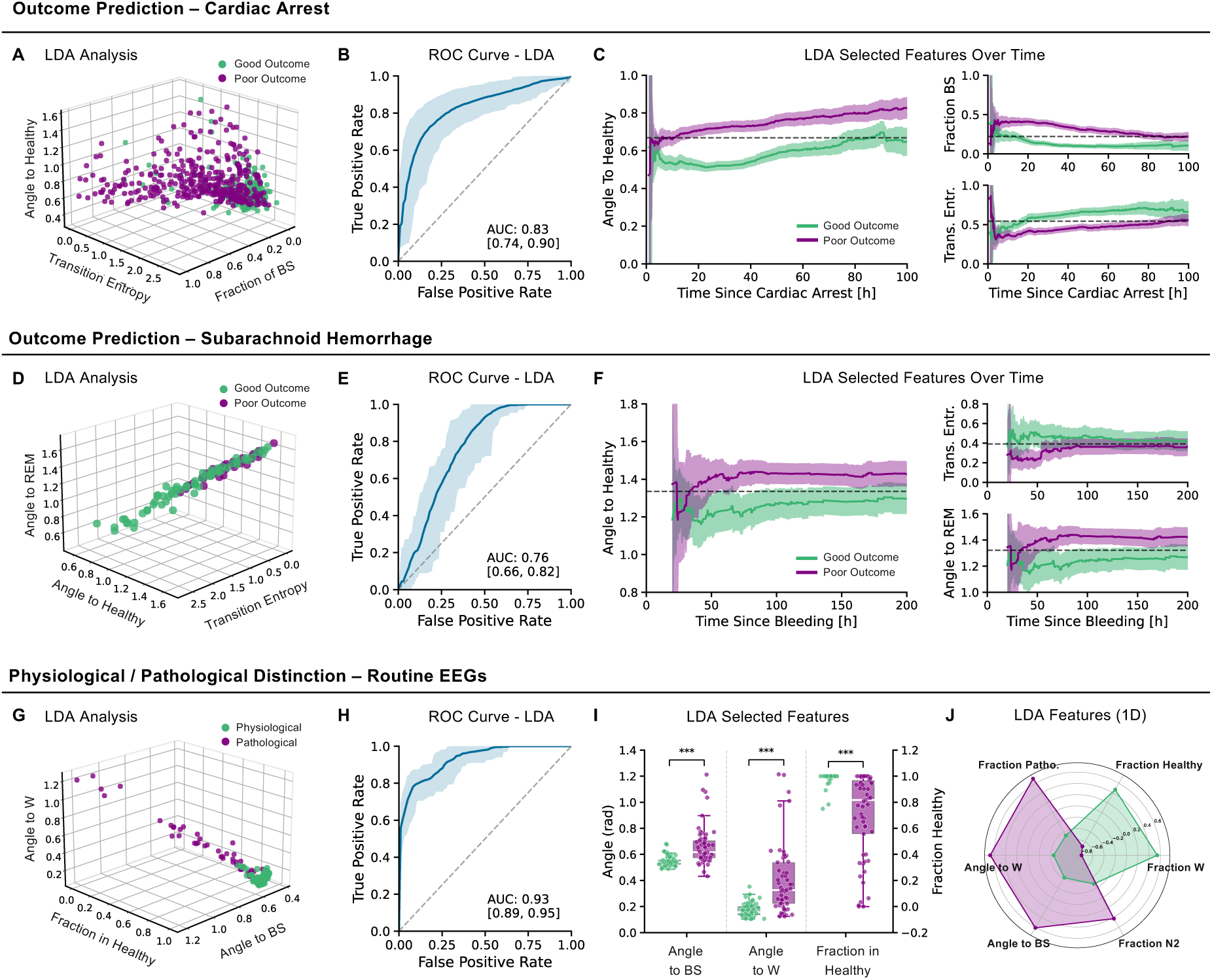
Trajectory feature analysis for outcome prediction across three cohorts. Panels **A–C** adapted from *Krumm et al.*, 2025 (Annals of Neurology) under CC BY 4.0 license. ROC curves were obtained by fitting an LDA hyperplane in the three-feature space; 95% confidence intervals were estimated via bootstrap resampling with 50 iterations. **A** 3D scatter plot of the three best LDA features for cardiac arrest patients with good (green) and poor (purple) outcome patients. **B** ROC curve for the LDA classifier of the CA cohort. **C** Cumulative running averages of the three LDA features over time since cardiac arrest, averaged across patients with 95% confidence intervals. Dashed horizontal lines indicate the midpoint between group means. **D** 3D scatter plot of the three best LDA features for SAH patients **E** ROC curve for the LDA classifier of the SAH cohort. **F** Cumulative running averages of the three LDA features over time since hemorrhage. **G** 3D scatter plot of the three best LDA features for routine EEG patients, with physiological (green) and pathological (purple) recordings. **H** ROC curve for the LDA classifier of the routine EEG cohort. I Box plots showing distributions of the three LDA features for physiological and pathological recordings. Significance was assessed using Mann-Whitney U tests (*** p<0.001). **J** Radar plot of the top six features ranked by individual discriminative ability, quantified by AUC of the raw feature value as decision score.

For long-term EEG in the SAH cohort, following the approach used for the CA data, features were also computed within non-overlapping 1-hour windows.^4^ This windowed analysis enables assessment of how features evolve over time, for example, whether the proportion of pathological states visited changes during the hospital stay. The three best features identified via LDA were plotted as cumulative running averages over time since onset of SAH, averaged across patients, with shaded 95% confidence intervals (Figure 2F).

For the routine EEG cohort, features were computed over entire recordings given their short duration (under 30 minutes). Group differences for the three LDA-selected features were assessed using Mann-Whitney U tests and visualized as box plots (Figure 2I). Additionally, the top six features were selected based on their individual ability to discriminate pathological from physiological EEGs, quantified by the AUC computed using the raw feature value as the decision score (Figure 2J).

#### Correlation of Trajectory Features and EEG Reports

To evaluate whether features derived from the 128D trajectories captured clinically relevant information contained in EEG reports beyond binary outcomes, we compared patient similarity based on UM-EEG-derived features to patient similarity derived from structured clinical reports. Specifically, for EEG-based similarity, z-scored features (Table 2) were compared pairwise across all patients using cosine similarity. For the report-based similarity, cosine similarity was computed for pairs of EEG reports from the z-scored 13 standardized report items (Table 1). The correlation between UM-EEG-derived and report-derived pairwise similarities was quantified using the Spearman correlation coefficient (Figure 3D).

**Figure 3.**
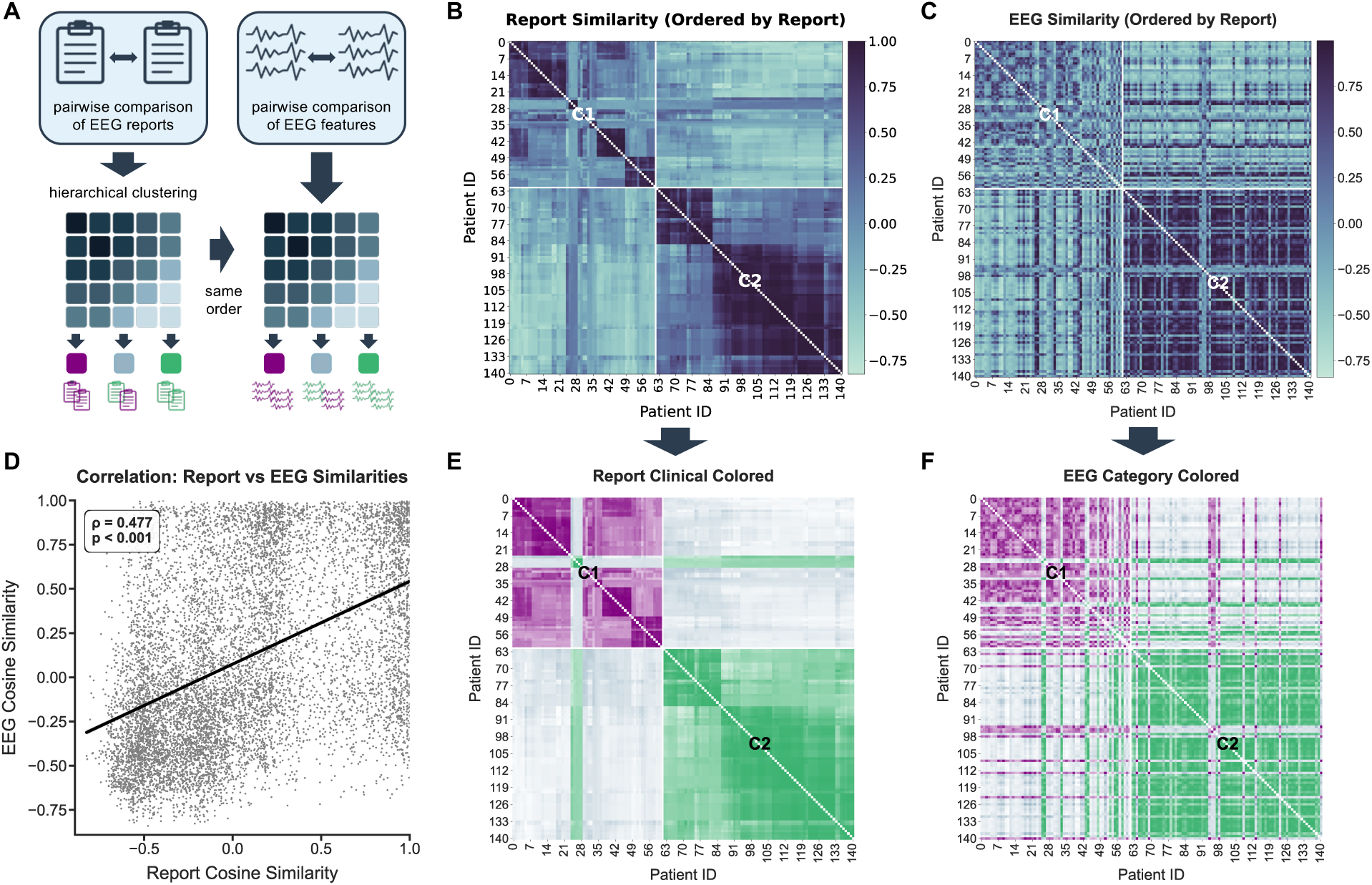
Comparison of report-based and EEG-based patient similarity. **A** Schematic of the approach: pairwise cosine similarities are computed separately from structured clinical report items and from EEG trajectory features. The resulting matrices are compared. Patient pairs are colored green if both patients are physiological, purple if both are pathological, and light blue if the two patients differ. For the report matrix, physiological or pathological labels are taken from clinical annotation (Items 9 and 10, Table 1). For the EEG matrix, labels are assigned by the LDA classifier trained on the six most discriminative EEG features (Fig.2J). **B** Patient-by-patient similarity matrix based on 13 structured report items, with patients ordered by hierarchical clustering (Ward’s method). Darker colors indicate higher cosine similarity. White lines mark the two cluster boundaries (C1, C2). The same matrix is shown below (**E**) colored by clinical annotation. **C** Patient-by-patient similarity matrix based on EEG trajectory features, with patients ordered identically to panel **B**. The same matrix is shown below (**F**) colored by LDA classifier prediction. The recovery of the report-derived cluster structure in the EEG matrix indicates that trajectory features capture groupings consistent with clinical assessment. **D** Scatter plot of all pairwise similarities. The significant positive correlation (Spearman ρ = 0.477, p < 0.001) indicates that patients judged similar by their clinical reports also show similar EEG trajectory patterns.

Additionally, both similarities were displayed as patient-by-patient similarity matrices. To visualize structure within the patient population, hierarchical clustering using Ward’s method was applied to the report-derived similarity matrix, constraining the solution to two clusters. The resulting patient ordering was then applied identically to the EEG-derived similarity matrix to allow direct visual comparison (Figure 3B, C).

To better illustrate the relationship between similarity structure and clinical labels, both matrices were coloured by outcome group: green if both patients were physiological, purple if both were pathological, and grey if the two patients had different labels. For the report matrix, labels were taken directly from the clinical annotation (items 9 and 10 in Table 1). For the EEG matrix, labels were assigned using a weighted composite score of the six most discriminative features. Discriminative ability of each feature was quantified by its AUC, computed by treating the raw feature value directly as a decision score. This means sweeping a threshold across all values and measuring how well each feature alone separates physiological from pathological recordings. Features were z-scored and weighted by their AUC before summation. Patients exceeding the optimal composite threshold were classified as pathological. To quantify the correspondence between the two-coloured matrices, pair-level colour agreement was computed across all patient pairs and summarized using Cohen’s κ.

### Interactive Visualization of Patient Trajectories in UM-EEG

To demonstrate how UM-EEG could serve as a tool for EEG contextualization and explainability, we developed a prototype web-based graphical user interface (GUI) for interactive exploration of EEG embeddings and patient trajectories within the UM-EEG coordinate space. We demonstrate the tool using routine EEGs as an example, given the availability of structured reports, though long-term EEG recordings can equally be displayed and summarized. The application was built using React for the frontend and Python for the backend.

The interface consists of four main components. First, a scatter plot displays the two-dimensional UMAP projection of the UM-EEG reference dataset. When a patient is selected, their trajectory is overlaid as a connected path and a pie chart summarizing the distribution of predicted EEG states across the recording is shown alongside. Second, a ‘Scored Reports’ panel provides an overview of each patient’s categorized report based on the 13 items in Table 1. Third, a segment viewer displays the raw four-channel EEG waveform (F3-C3, C3-O1, F4-C4, C4-O2) for any selected time point, with navigation via a slider or by clicking directly on trajectory points. Fourth, a ‘Similar Patients’ panel identifies the three most similar patients based on cosine similarity of features extracted from the 128D embedding space, displaying their trajectories alongside their associated clinical reports to enable rapid comparison with reference cases.

### Statistical Testing

Continuous variables, such as recording durations, are summarized as median and interquartile range (IQR), where IQR denotes the range from the 25th to the 75th percentile. To compare how much time patients with different outcomes spent in each of the 11 UM-EEG reference states, we used two-sided Mann-Whitney U tests, applied separately for each state within each cohort. The same test was used to compare the three LDA-selected features between physiological and pathological recordings in the routine EEG cohort. To quantify how well the structure of the EEG-derived patient similarity matrix matched the structure of the report-derived similarity matrix, we used the Spearman rank correlation coefficient on all pairwise similarity values. Agreement between the two matrices at the level of individual patient pairs was quantified using Cohen’s κ. A significance threshold of p < 0.05 was applied throughout.

## Results

### Datasets and embedding into a shared reference space

We applied UM-EEG to three datasets spanning distinct clinical contexts. The cardiac arrest (CA) cohort (n=576) comprised long-term EEG recordings from patients with disorders of consciousness in the ICU. The second dataset, subarachnoid hemorrhage (SAH) patients (n=100), represented a similar critical care setting with prolonged EEG monitoring. The third, routine EEGs (n=141), spanned both healthy and pathological patterns in a short-term recording context, extending the framework to a broader and more heterogeneous clinical use case. For each dataset, patient recordings were mapped into the shared 128D embedding space and visualized as trajectories relative to the 11 UM-EEG reference states.

The three cohorts differed substantially in recording duration, continuity and coverage, reflecting their distinct clinical contexts (Figure 1A–F). CA recordings were relatively consistent, with a median actual EEG duration of 1.87 days (IQR: 1.08–2.76) spanning a median monitoring window of 2.65 days (IQR: 1.88–3.67), and comparable coverage across outcome groups. SAH recordings, while spanning a considerably longer monitoring window (median 5.93 days, IQR: 3.13–8.99), contained far less usable EEG data: the median actual recording duration was only 0.55 days (IQR: 0.27–1.73), indicating that monitoring was frequently interrupted. Reconstruction of 10-second epochs from 5-second snippets resulted in a mean data loss of 22.6% (95% CI: 17.8–27.4%) due to exclusion of non-consecutive or artifactual segments. Recording coverage also varied substantially between patients, with 46% having less than half a day of EEG data and a coefficient of variation of 1.41, roughly twice that of the CA cohort (0.72). Routine EEG recordings were short and continuous, with a median duration of 20 minutes (IQR: 18–20 min).

Figure 1G shows the 2D projection of all three cohorts into the shared UM-EEG embedding space. The visualization conveyed qualitative differences between outcome groups while statistical comparisons were performed in 128D space. As shown in Krumm et al.,^4^ the 2D projection closely preserves the spatial arrangement of the 128D space, verified via multidimensional scaling of class medians and their cosine distances.

### Outcome-dependent occupancy of embedding states

Across all three cohorts, a consistent pattern emerged: patients with good outcome spent significantly more time in healthy embedding states than those with poor outcome (CA: 32.5% vs 27.4%, p<0.001; SAH: 8.9% vs 1.3%, p=0.002; routine EEG: 96.4% vs 70.2%, p<0.001). In CA patients, burst suppression showed the most pronounced difference, with more than threefold higher occupancy in patients with poor outcome (30.9% vs 8.9%, p<0.001). Good outcome patients showed substantially more time in sleep stages, particularly REM (13.4% vs 5.5%, p<0.001) and N1 (10.1% vs 5.3%, p<0.001), consistent with preservation of sleep cycling as a marker of cortical recovery.^17,18^ In SAH patients, both groups (good and poor outcome) spent the majority of time in pathological states, reflecting the severity of the acute injury. Again, good outcome patients nonetheless spent more time in sleep stages including N1 (p<0.001), N2 (p<0.001), N3 (p=0.011), and REM (p=0.001), with overall healthy state occupancy nearly sevenfold higher (8.9% vs 1.3%). Among pathological states, only GRDA significantly differentiated the groups, being higher in poor outcome patients (29.3% vs 18.7%, p=0.007). In routine EEGs, physiological recordings were concentrated in wakefulness (81.7% vs 39.2%, p<0.001), while pathological recordings showed markedly increased LRDA (10.1% vs 0.2%), GRDA (10.7% vs 0.8%), and SZ (3.1% vs 0.5%; all p<0.001).

### Trajectory-derived features enable outcome discrimination

We next investigated whether trajectory-based features derived from UM-EEG could distinguish clinical outcomes and physiological states across cohorts. Consistent with Krumm et al., UM-EEG trajectory features effectively discriminated outcomes in cardiac arrest patients, with burst suppression occupancy, distance to the healthy centroid, and state transition entropy identified as the three most informative features (Figure 2A-C; ROC-AUC 0.83, 95% CI: 0.74-0.90). In SAH patients, brute-force LDA selected angle to REM, angle to the healthy centroid, and state transition entropy as the optimal three-feature combination (Figure 2D-F), achieving a ROC-AUC of 0.76 (95% CI: 0.66-0.82). When plotted as cumulative running averages over time since bleeding, angle to REM and angle to the healthy centroid consistently maintained non-overlapping confidence intervals between outcome groups for most of the monitoring period, with overlap occurring only during the early hours when data were sparse. In contrast, state transition entropy exhibited partial overlap throughout, providing less stable separation over time.

For routine EEGs, the optimal features were angle to the W state, fraction of time in healthy states, and angle to BS (Figure 2G-I), yielding a ROC-AUC of 0.93 (95% CI: 0.89–0.95). All three features differed significantly between physiological and pathological recordings (all p<0.001). Physiological recordings clustered tightly around the wakefulness state, producing a consistent angular distance to BS. Pathological recordings spread more broadly across the embedding space, increasing both the mean and variance of the angle to BS. The feature therefore captured heterogeneity in brain state as displacement within the embedding space rather than reflecting burst suppression occupancy per se. The top six features ranked by individual discriminative ability were fraction of time in pathological states, fraction in healthy states, fraction in W, angle to W, angle to BS, and fraction in N2 (Figure 2J). Together, these results demonstrate that UM-EEG trajectory features capture robust, geometry-based signatures of brain state that discriminate clinical outcomes and physiological versus pathological patterns across diverse EEG cohorts.

### Embedding geometry recapitulates clinical report structure

To assess whether UM-EEG trajectory features capture clinically meaningful information consistent with that contained in structured clinical EEG reports, we compared patient similarity derived from EEG features to similarity derived from report items (Figure 3). Hierarchical clustering of the report-based similarity matrix revealed two dominant clusters, C1 and C2, broadly corresponding to pathological and physiological patient groups respectively (Figure 3B, E). Remarkably, when the EEG-based similarity matrix was ordered identically, the same two clusters were clearly recoverable, indicating that EEG trajectory features alone reproduce the principal grouping structure found in clinical reports, without access to report labels (Figure 3C, F).

This correspondence was confirmed quantitatively at the level of individual patient pairs. Across all pairs, EEG-derived and report-derived similarity matrices showed substantial agreement (73.4% exact match, κ = 0.570). Restricting to pairs with unambiguous labels (both patients either physiological or pathological) yielded near-perfect agreement (97.2%, κ = 0.936). At the overall similarity level, pairwise EEG cosine similarity and report cosine similarity were significantly positively correlated (Spearman rank correlation ρ = 0.477, p < 0.001; Figure 3D), demonstrating that patients judged similar by expert reports also exhibited similar EEG trajectory patterns in the 128D embedding space (Figure 3D). Across cohorts, these findings show that UM-EEG captures a consistent, geometry-based organization of brain states that aligns closely with expert interpretation and generalizes across diverse clinical contexts.

### Interactive visualization of patient trajectories

To support interpretation and clinical integration of UM-EEG embeddings, we developed an interactive graphical user interface (GUI) for exploring patient trajectories (Figure 4). The main view displays a 2D UMAP projection of the reference dataset, with points color-coded by EEG pattern and the selected patient’s trajectory overlaid with a temporal gradient (Figure 4A). A detail panel summarizes state distribution via a pie chart (Figure 4B), while a segment viewer allows inspection of raw EEG at specific time points (Figure 4C). Similar trajectories from other patients can be retrieved using cosine similarity in the 128D space, enabling side-by-side comparison with associated clinical reports (Figure 4D). This interactive tool provides structured, intuitive EEG interpretation and illustrates how deep embedding-based representations can be integrated into clinical workflows, helping to close the translational gap between computational analyses and bedside decision-making.

**Figure 4.**
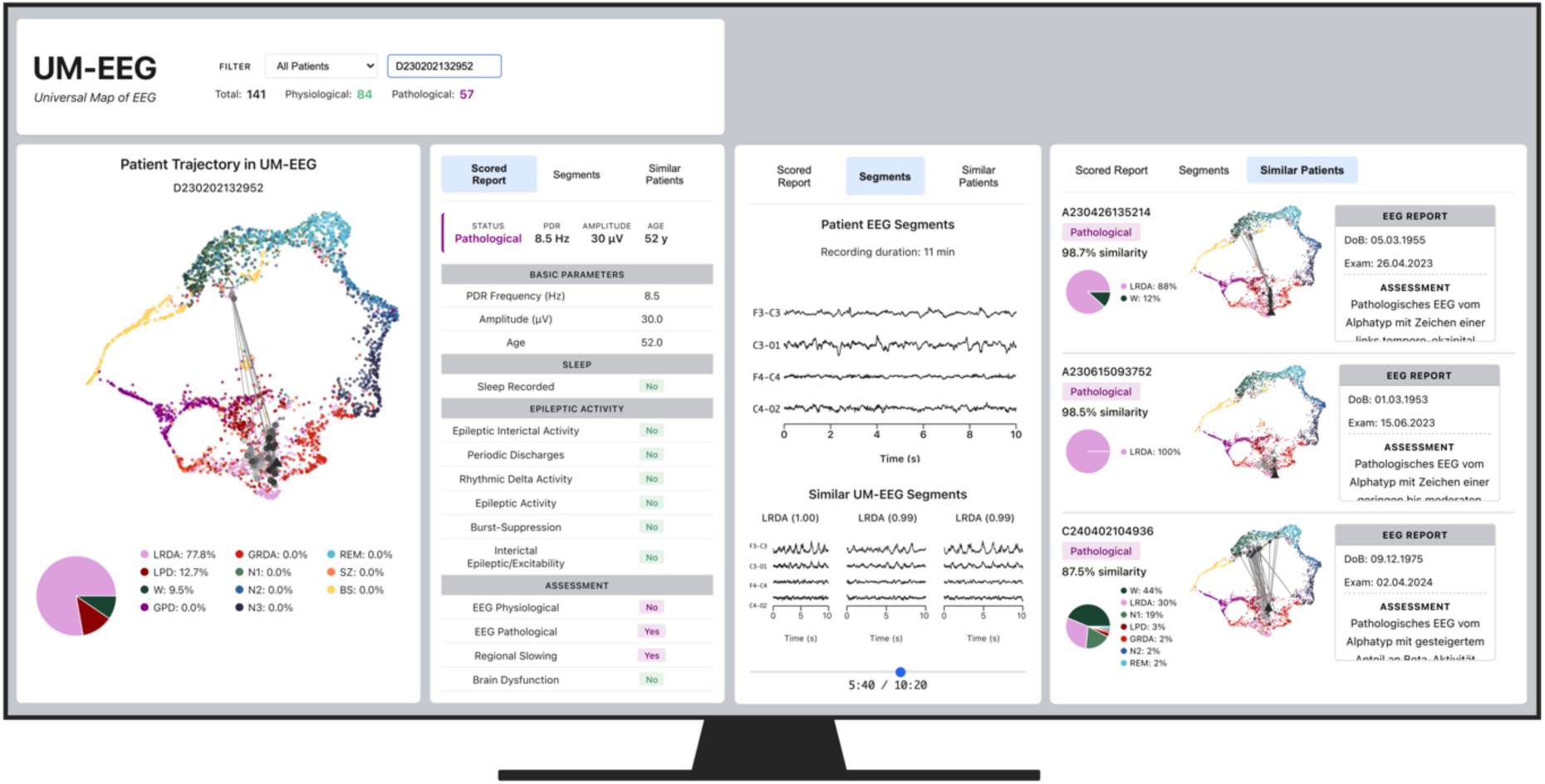
An interactive tool for simpler EEG evaluation. **A** Two-dimensional UMAP projection of the UMEEG reference dataset, with points color-coded by EEG pattern. The selected patient’s trajectory is overlaid as a connected path with temporal gradient (white: recording start; black: recording end). Triangle markers indicate trajectory boundaries. **B** Trajectory detail panel showing the patient’s path through embedding space with corresponding pie chart summarizing the distribution of predicted EEG states across the recording. **C** Segment viewer displaying the raw four-channel EEG waveform (F3-C3, C3-O1, F4-C4, C4-O2) for the selected time point. Users can navigate segments via slider or by clicking trajectory points. **D** Similar trajectories panel showing the three most similar patients identified by cosine similarity in 128-dimensional embedding space, with their trajectories and associated clinical reports displayed for comparison.

## Discussion

UM-EEG is founded on a simple but powerful idea: train a latent embedding space on expert-defined EEG patterns (wake, sleep stages, burst suppression, and ictal-interictal continuum patterns) and use it as a fixed reference map into which any new or ambiguous recording can be projected and interpreted. This approach rests on the assumption that brain states exist along a low-dimensional continuum,^19^ and that a semantic, continuous reference space places novel data relative to well-characterized patterns.

Our results highlight four key aspects. First, the pretrained space generalizes across new datasets and medical conditions, including subarachnoid hemorrhage (SAH) and routine clinical EEGs, which differ from the original cardiac arrest (CA) cohort in etiology, monitoring duration, and clinical setting. Second, while trajectory features predict outcome, the more clinically meaningful contribution may be providing a framework for EEG interpretation that bridges the gap between oversimplified binary scores and raw, unwieldy long-term recordings. Third, a prototype GUI illustrates how embedding-based trajectories can be integrated into clinical workflows, contextualizing EEG evolution over time and enabling comparison to similar patients. Finally, we discuss limitations and future directions to enhance the fidelity and applicability of the embedding.

The two novel datasets test generalizability in complementary ways. SAH patients, monitored continuously in the ICU, present with hemorrhagic injury and distinct recovery dynamics, while routine EEGs represent short, outpatient recordings covering both physiological and pathological patterns. Importantly, neither dataset was included in training, and the UM-EEG was applied without retraining or adjustment.

Trajectory-derived features yielded meaningful separation across all cohorts. In SAH, outcome discrimination achieved an AUC of 0.76, while routine EEGs distinguished physiological and pathological recordings with an AUC of 0.93, using only three features derived from proximity reference anchors. Beyond predictive performance, the features themselves reveal pathophysiologically interpretable patterns. In CA, burst suppression dominated, consistent with the existing literature. In SAH, proximity to REM and distance from the healthy centroid were most predictive, reflecting the prognostic relevance of preserved sleep in disorders of consciousness.^18,20^ In routine EEGs, proximity to wakefulness was most discriminative, while distance from burst suppression captured displacement from the healthy cluster rather than true pathological occupancy. These observations highlight both the strengths and limitations of current geometric features: they capture relative positioning but do not always distinguish between true occupancy and displacement. Nevertheless, a consistent principle emerges across cohorts: the closer a patient’s EEG to healthy reference states, the better the outcome.

The prototype GUI demonstrates how such embedding could support bedside interpretation. Clinicians can visualize a patient’s trajectory relative to the reference apace, inspect raw EEG segments at any point, and retrieve similar patient trajectories with corresponding reports for direct comparison. Unlike existing EEG tools focused on seizure detection,^21–23^ alerting or quantitative trending, UM-EEG provides contextualization: summarizing temporal evolution, anchoring recordings to established brain states, and enabling structured comparison across patients. Integration into clinical workflows could allow real-time monitoring of deviation from healthy states or transitions into pathological patterns, helping close the translational gap between computational models and bedside decision-making.

Several limitations warrant discussion. First, reducing trajectories to distances from reference states necessarily discards information about the full geometry of the recording. Future work could leverage the full 128D coordinates while preserving interpretability. Second, the current encoder uses only four bipolar channels and spectrogram representations, which may omit relevant signal features. Transformer-based models operating on raw EEG and handling variable channel configurations could improve fidelity. Third, finer-grained events, such as epileptiform spikes, were not included in the training set, leaving open questions about where such patterns would fall within the embedding. Nevertheless, sub-structure already visible within clusters, such as ordering by suppression ratio in the burst suppression cluster, suggests the latent space captures more nuance than the reference labels alone convey.

In summary, UM-EEG demonstrates that semantic, continuous latent spaces can provide interpretable, clinically relevant EEG representations. The deviation from healthy brain states over time offers a simple yet powerful summary, capturing prognostic information across diverse patient populations and recording contexts. Reference states act as anchors, maintaining interpretability without forcing recordings into rigid categories, echoing the contextual reasoning clinicians use when reading EEG. By bridging the gap between complex, high-dimensional recordings and actionable clinical insights, embedding-based approaches offer a pathway toward scalable, generalizable, and clinically integrated EEG interpretation.

## Data Availability

Most data produced are available online at https://bdsp.io. The Charite dates is available upon reasonable request to the authors.

## Acknowledgements

Dr. Westover was supported by grants from the NIH (RF1AG064312, RF1NS120947, R01AG073410, R01HL161253, R01NS126282, R01AG073598, R01NS131347, R01NS130119), and NSF (2014431). Dr. Meisel acknowledges support from the Schilling Foundation Professorship Computational Neurology.

## Author Contributions

C.M. and L.K. contributed to the conception and design of the study. R.T. and M.B.W. contributed to the acquisition of data. L.K. prepared the figures and drafted the text. L.K. pre-processed and analysed the data and applied all machine learning methods. D.K. prepared trajectory features. C.M. and L.K. wrote the manuscript, all authors edited the manuscript.

## Data Sharing

Data supporting the study is partly publicly accessible. This includes the Cardiac Arrest data https://doi.org/10.60508/xwye-v214 and the Subarachnoid Hemorrhage data https://bdsp.io/content/saheeg/1.0/. The routine EEG data is available upon reasonable request.

## Code Availability

Code is available at https://gitlab.com/computational-neurologie/.

